# Exploring the relationship between women’s experience of postnatal care and reported staffing measures: an observational study

**DOI:** 10.1101/2022.04.03.22272935

**Authors:** Lesley Turner, Jane Ball, David Culliford, Ellen Kitson-Reynolds, Peter Griffiths

**Author notes:** corresponding author (LT).

## Abstract

**Background:** Women have reported dissatisfaction with care received on postnatal wards and this area has been highlighted for improvement. Studies have shown an association between midwifery staffing levels and postnatal care experiences, but so far, the influence of registered and support staff deployed in postnatal wards has not been studied. This work is timely as the number of support workers has increased in the workforce and there has been little research on skill mix to date.

**Methods:** Cross sectional secondary analysis including 13,264 women from 123 postnatal wards within 93 hospital Trusts. Staffing was measured at organisational level as Full Time Equivalent staff, and at ward level using Care Hours Per Patient Day. Women’s experiences were assessed using four items from the 2019 national maternity survey. Multilevel logistic regression models were used to examine relationships and adjust for maternal age, parity, type of birth, medical staff and number of births per year in the Trust.

**Results:** Trusts with higher levels of midwifery staffing had higher rates of women reporting positive experiences of postnatal care. However, when staffing was measured at a ward level, there was no evidence of an association between registered staffing and patient experience. Wards with higher levels of support worker staffing were associated with higher rates of women reporting they had help when they needed it and were treated with kindness and understanding.

**Conclusion:** The relationship between reported registered staffing levels on postnatal wards and women’s experience is uncertain. Further work should be carried out to examine why relationships observed at an organisational level were not replicated closer to the patient, at ward level. It is possible that reported staffing levels do not reflect staff as deployed if midwives are floated to cover delivery units. This study highlights the potential contribution of support workers in providing quality care on postnatal wards.

## Background

Midwives have expressed concern about the quality of postnatal care provided in hospital, and cite increased workloads, limited staffing and busy work environments as contributing to this[1, 2]. In line with this, women have reported lower satisfaction with postnatal care compared with antenatal and labour care[3, 4]. A survey of 1260 first time mothers found that less than half of them had all the help they needed with infant feeding, one in five did not have the physical care they needed, and one in seven had unmet information needs[5].

The quality of care is affected by the availability of staff[6] and is it reported that staff prioritise those with urgent medical needs and consider discharging other women to ease pressure on beds[7]. Staff need to be available with the right skills, in the right place at the right time to deliver high quality care and minimise avoidable harm[8]. Staffing shortages in postnatal care have been highlighted in a number of reports[9-11] and have been linked with a lack of support for breastfeeding, delays in care and dissatisfaction for families. Studies on satisfaction with postnatal care are now emerging, highlighting the need for individualised responsive care[12], proactive information and emotional support[13]. It had previously been suggested that dissatisfaction with postnatal care was related to a mismatch between expectations and experiences, however recent work by McLeish et al [12] has disputed this and highlights the meeting of individual needs as the key issue.

The average length of postnatal stay has reduced from 2.8 days in 2001 to 1.5 days in 2020[14], and almost half of women are discharged within 24 hours[15]. Faster patient turnover leads to a greater proportion of the midwives’ day involved in admissions or discharges, which reduces the amount of time spent providing ongoing care[14]. Reduced length of stay means that people staying in wards may be more acutely unwell than before[16]. Increased risk factors such as rising maternal age, obesity, diabetes and rates of intervention have contributed to rising acuity in postnatal women[17-19].

This change in patient population and ward activity needs to be reflected in the planned staffing to ensure a safe and effective service. From a workforce perspective, it is not clear whether the deployed workforce has kept up with demand and acuity. Variability can be seen across England in the number of midwives and health care support workers present on postnatal wards[20]. The development of support workers and growth in their numbers has led to expansion of these roles on many postnatal wards[21] and it is uncertain whether this growth is based on evidence of benefit.

One cross sectional study found that higher registered midwife workforce is associated with a better postnatal care experience, as measured by the National Maternity Survey in England[22]. This study measured staffing at an organisational level across the whole maternity service. We found no published studies examining the impact of staffing measured at a postnatal ward level, which is expected to provide a more accurate measurement of staffing exposure. A study by Winter et al[23] found that aggregation of data changes the magnitude of estimation when examining the association between staffing and outcomes. Point estimates for associations were larger in magnitude when more precise data on staffing was used.

We set out to use staffing at a ward level to improve estimates of exposure and to include the contribution of support workers as this is a growing workforce that has received little attention in research studies to date. Failure to control for differences in support worker staffing may distort relationships between midwife staffing and patient experiences, or may lead to an overemphasis on registered staff to address quality of care problems[24].

Since 2018 staffing on all acute hospital wards in England has been made available in the Care Hours Per Patient Day dataset (CHPPD)[20]. This new data source presents an opportunity to explore ward level staffing in closer proximity to patient experience, and to include support worker staffing in regression models. Although Nursing Hours per Patient Day has been measured in many primary studies[25, 26], the dataset of CHPPD[20] has not been used in published research to our knowledge so its contribution and limitations have yet to be described.

In this paper we seek to explore the relationship between staffing levels in midwifery services and women’s experiences of postnatal care in inpatient wards. The following research questions were studied :

- Is there an association between midwife staffing in the organisation and women’s experience of postnatal care?
- Is there an association between postnatal ward staffing levels and composition (registered nurses, midwives and support workers) and the experience of mothers receiving postnatal care?

## Methods

This is a cross sectional analysis of linked routinely collected datasets in English hospital Trusts. Anonymised individual patient data from the 2019 National Maternity Survey was obtained from the UK Data Service[3]. This data relates to women’s experience of maternity care and the survey was sent to all women who had a live birth in February 2019 under the care of an NHS Trust. Case mix variables of age band, parity, and type of birth were extracted for individuals participating in the survey. Length of stay was not extracted as it could be both a measure of case mix or outcome measure and is also likely to be highly correlated with type of birth. Ethnicity and response rate per Trust were obtained from data published online relating to this survey. This study was reviewed and approved by the University of Southampton Ethics and Research Committee prior to data collection (ERGO 62570). All data was fully anonymized before they were supplied for this secondary analysis. Staffing data was obtained at a ward level via the Care Hours Per Patient Day (CHPPD) dataset which is publicly available on NHS England website[20]. CHPPD is available for Registered staff (combined for Nurses or Midwives) and separately for Health care support staff. This is reported as a monthly figure for each staff group in the dataset. CHPPD is calculated from the number of patients occupying beds at midnight each day and the actual hours worked by staff groups, totalled for the month. These totals are then divided to produce the CHPPD average for that month[27]. Hours worked by permanent, bank and agency staff are included, but those worked by supernumerary staff and students are not. Newborns were not counted in addition to mothers in this dataset, although this measure has been introduced for subsequent CHPPD datasets from 2021.

Workforce data from the single month of February 2019 was used as this matches the period of the patient experience survey. Postnatal wards were identified by selecting the speciality codes of Midwifery or Obstetrics within the dataset. Entries were excluded if they were unlikely to be postnatal wards e.g. if Registered staff CHPPD >=10 or if they were named exclusively as Labour Ward, Delivery Suite, Birth centre, Antenatal ward or Neonatal unit. The 10 CHPPD cut off was chosen on examination of the CHPPD data by ward title in the whole dataset, where it emerged that CHPPD records above this level were found in critical care areas and labour wards, and this is supported by the categorisation by Twigg and Duffield [28]. This process identified 94 Trusts containing 124 wards. 27 Trusts were not represented as postnatal wards were not identifiable using the method above. Fifteen Trusts had more than one postnatal ward, so the published February 2019 CHPPD for each ward was averaged to produce one figure per Trust for each staff group. The variable of skill mix was generated by calculating the percentage of registered staff on each ward.

Additional analyses of midwifery staffing across services were undertaken using staffing measured at an organisational (i.e. Trust) level as in our earlier study[22]. This newly obtained 2019 dataset was used to confirm or counter our previous findings at a Trust level. We included Full Time Equivalent midwives and medical staff employed in Obstetric and Gynaecology[29] although support worker figures could not be ascertained at this level. Workload for each staff group was measured by dividing the number of staff by the number of births per year in the Hospital Episode Statistics dataset [30].

Four survey questions were selected for analysis (Table 1) as they were the only questions which explored the quality of care provided by staff on the postnatal ward. These questions were answered only by women who received care in hospital after the birth due to filter questions within the survey.

**Table 1.** Questions in maternity survey selected for analysis in this study

| Questions in Maternity Survey | Response options |
| --- | --- |
| On the day you left hospital, was your discharge delayed for any reason? | Yes<br>No |
| If you needed attention while you were in hospital after the birth, were you able to get a member of staff to help you when you needed it? | Yes, always<br>Yes, sometimes<br>No<br>I did not want/need this<br>Don't know/can't remember |
| Thinking about the care you received in hospital after the birth of your baby, were you given the information or explanations you needed? | Yes, always<br>Yes, sometimes<br>No<br>Don't know/can't remember |
| Thinking about the care you received in hospital after the birth of your baby, were you treated with kindness and understanding? | Yes, always<br>Yes, sometimes<br>No<br>Don't know/can't remember |

The survey ordinal responses were dichotomised into ‘yes always’=1 and ‘yes sometimes or no’ = 0 based on an implied quality standard[31]. Alternative grouping was tested in the sensitivity analysis. ‘Don’t know/can’t remember’ responses represented less than 1% of the data for each question and were removed by pairwise deletion to maximise the data available for analysis. The patient experience variables were linked to staffing variables by the unique Trust code for each organisation.

We were able to link ward level CHPPD data to patient experience in 93 Trusts with 123 wards, representing 13,264 respondents. Trusts were divided into tertiles to represent those with the highest, middle and lowest staffing for both registered and support staff. The rationale was to enable detection of non-linear effects which would not be possible if analysed as continuous variables, and it also aids interpretation[32]. A secondary analysis with staffing as a continuous variable was also conducted.

Descriptive analysis was performed to understand the variation in staffing between postnatal wards and between Trusts. Variation in women’s responses between Trusts were summarised. A two-level multilevel logistic regression model was created using Level-1 (mothers) nested within Level-2 (Trusts). The null model was a two-level random intercept model with no predictors to explore the extent of between-trust variation in the outcomes. Covariates of mothers’ age band, parity, type of birth and midwifery staffing measures were included in the main model as these characteristics have been shown to contribute to the greatest amount of variability in clinical outcomes and patient experience measures[22, 33]. Additional covariates of survey response rate, ethnic case mix of Trust, number of births in each Trust and obstetric/gynaecological medical staffing were added to the main model and were retained only if the model fit improved. These variables were added as it was anticipated that they could account for variation in women’s satisfaction or because they may highlight bias in the self-selection of respondents.

Model fit was judged by calculating the Akaike’s Information criterion (AIC) and Bayesian information criterion (BIC). If AIC and BIC scores disagreed, then priority was given to the model lowest on AIC, and the model lower on BIC was scrutinised and compared for a sensitivity check (see supplementary material S1 for details of variable selection and model fit). The primary model considered Registered staff and Support workers as independent groups in the workforce. It is also possible to conceptualise the workforce as a single entity with varying composition, and so alternative models with variables representing skill mix (percentage registered staff) and total number of staffing hours (combined CHPPD for registered and support staff) were explored as secondary analyses.

## Results

The characteristics of respondents in the maternity survey are given in Table 2. The median response rate was 39% among Trusts (IQR 33% to 42%).

**Table 2:** Data characteristics for respondents in Maternity Survey

|  | Level of data | Summary<br>Medians presented due to skewed distributions |
| --- | --- | --- |
| Response rate | Trust level | Median 38.7%<br>IQR 32.6%, 42.4% |
| Age group of mothers | Individual patient | 16-25 years 6.6%<br>25-29 years 20.0%<br>30-34 years 37.5%<br>35+ years 35.9% |
| Parity | Individual patient | Primiparous 50.9%<br>Multiparous 49.1% |
| Type of birth | Individual patient | Spontaneous birth 55.4%<br>Instrumental birth 14.7%<br>Planned caesarean birth 13.8%<br>Emergency caesarean birth 16.1% |
| Percentage white ethnicity | Trust level | Median 84.7%<br>IQR 73.5%, 91.2% |

At organisational level the median FTE midwives per 100 births was 3.58 (IQR 3.33 to 3.84), equivalent to one midwife for every 28 births. For the 123 postnatal wards included in the analysis, the median CHPPD for registered staff on postnatal wards was 4.69 (IQR 3.75, 5.80) and for support workers it was 2.46 (IQR 1.91, 3.18) (Table 3). The median percentage of registered staff on postnatal wards was 63.6% (IQR 58.0%, 70.6%).

**Table 3:**
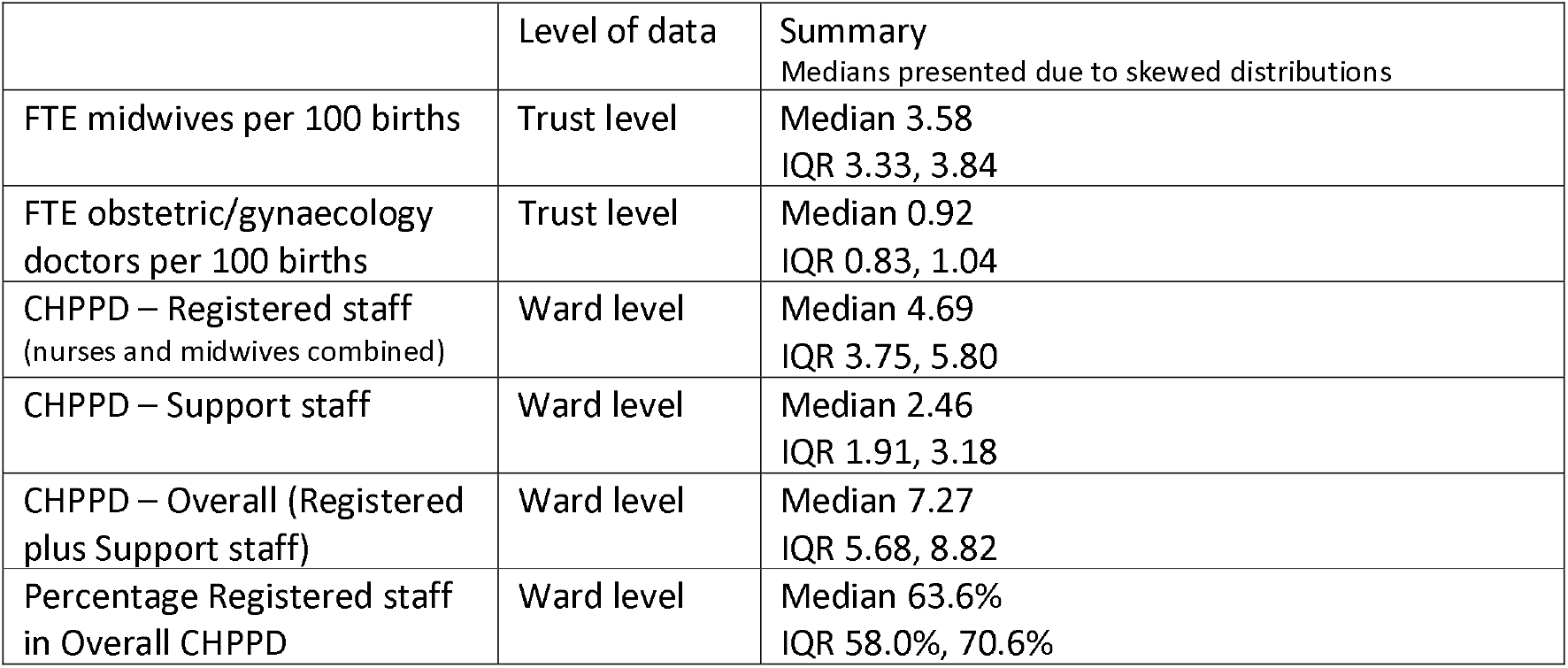
Distribution of staffing at ward level and organisation level

|  | Level of data | Summary<br>Medians presented due to skewed distributions |
| --- | --- | --- |
| FTE midwives per 100 births | Trust level | Median 3.58<br>IQR 3.33, 3.84 |
| FTE obstetric/gynaecology doctors per 100 births | Trust level | Median 0.92<br>IQR 0.83, 1.04 |
| CHPPD – Registered staff (nurses and midwives combined) | Ward level | Median 4.69<br>IQR 3.75, 5.80 |
| CHPPD – Support staff | Ward level | Median 2.46<br>IQR 1.91, 3.18 |
| CHPPD – Overall (Registered plus Support staff) | Ward level | Median 7.27<br>IQR 5.68, 8.82 |
| Percentage Registered staff in Overall CHPPD | Ward level | Median 63.6%<br>IQR 58.0%, 70.6% |

15 Trusts were in the highest tertile for both Registered CHPPD hours and Support staff CHPPD hours. 21 Trusts were in the lowest tertiles for both these staffing measures. Three Trusts were in the lowest tertile for Registered staffing while also having the highest tertile of support staff CHPPD. The Trust level measure of Registered FTE Midwives and the ward level measure of Registered staffing CHPPD appear to have only a weak relationship between them, Spearman’s rho 0.197 (see supplementary material S2 for more detail on staffing distribution at Trust and ward level.

### Summary of patient experience

Responses to the four questions relating to postnatal care are given in Table 4. The majority of women answering each question responded positively, although the worst rated measure was delay in discharge with 44% of women stating they had experienced a delay. 75% of respondents reported they were always treated with kindness and understanding.

**Table 4.** Summary of postnatal experience measures

| Question in Maternity Survey | Response categories | Frequency<br>% answers | Statistically sig<br>variables in UV<br>analysis |
| --- | --- | --- | --- |
| On the day you left hospital, was your discharge delayed for any reason?<br>n= 13,264, 450 missing | Yes<br>No | 5690 (44.4%)<br>7124 (55.6%) | Age group<br>Parity<br>Type birth |
| If you needed attention while you were in hospital after the birth, were you able to get a member of staff to help you when you needed it?<br>n=13,264, 420 missing<br>827 not applicable as did not want/need help | Yes, always<br>Yes, sometimes<br>No<br>Don't know/can't remember | 7376 (61.4%)<br>3829 (31.9%)<br>762 (6.3%)<br>50 (0.4%) | Parity<br>Type birth |
| Thinking about the care you received in hospital after the birth of your baby, were you given the information or explanations you needed?<br>n=13,264, 402 missing | Yes, always<br>Yes, sometimes<br>No<br>Don't know/can't remember | 8361 (65%)<br>3507 (27.3%)<br>890 (6.9%)<br>104 (0.8%) | Parity<br>Type birth |
| Thinking about the care you received in hospital after the birth of your baby, were you treated with kindness and understanding?<br>n=13,264, 400 missing | Yes, always<br>Yes, sometimes<br>No<br>Don't know/can't remember | 9653 (75%)<br>2771 (21.5%)<br>405 (3.2%)<br>35 (0.3%) | Age group<br>Parity<br>Type birth |

Univariable analyses using the dichotomised data for patient response found that age group, parity, type of birth were significantly associated with differences in response to some questions (see supplementary material S3).

### Relationship between staffing and patient experience

#### Trust Level

When analysed at a Trust level, higher staffing levels of Registered midwives were associated with better patient experience measures in terms of delay without discharge and women always receiving the information and explanations they needed. Findings remained statistically significant after controlling for case mix factors of age, parity and type of birth. Table 5 includes a summary of the direction of effects, the point estimates for odds ratios and confidence intervals. Women were more likely to report they always had help when needing it and been treated with kindness and understanding in Trusts with higher numbers of midwives, but these findings were not statistically significant.

**Table 5:**
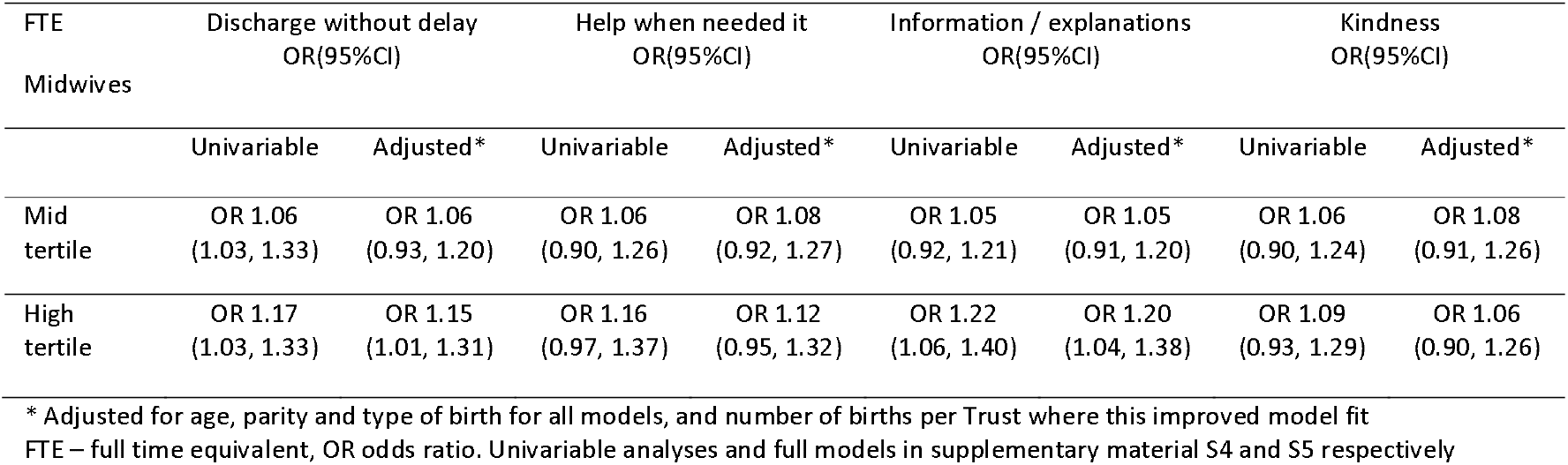
Summary of univariable and adjusted regression analysis. Estimated odds of a positive response compared to the lowest tertile of staffing (analysed at Trust level).

| FTE<br>Midwives | Discharge without delay<br>OR(95%CI) |  | Help when needed it<br>OR(95%CI) |  | Information / explanations<br>OR(95%CI) |  | Kindness<br>OR(95%CI) |  |
| --- | --- | --- | --- | --- | --- | --- | --- | --- |
|  | Univariable | Adjusted* | Univariable | Adjusted* | Univariable | Adjusted* | Univariable | Adjusted* |
| Mid<br>tertile | OR 1.06<br>(1.03, 1.33) | OR 1.06<br>(0.93, 1.20) | OR 1.06<br>(0.90, 1.26) | OR 1.08<br>(0.92, 1.27) | OR 1.05<br>(0.92, 1.21) | OR 1.05<br>(0.91, 1.20) | OR 1.06<br>(0.90, 1.24) | OR 1.08<br>(0.91, 1.26) |
| High<br>tertile | OR 1.17<br>(1.03, 1.33) | OR 1.15<br>(1.01, 1.31) | OR 1.16<br>(0.97, 1.37) | OR 1.12<br>(0.95, 1.32) | OR 1.22<br>(1.06, 1.40) | OR 1.20<br>(1.04, 1.38) | OR 1.09<br>(0.93, 1.29) | OR 1.06<br>(0.90, 1.26) |

\* Adjusted for age, parity and type of birth for all models, and number of births per Trust where this improved model fit FTE – full time equivalent, OR odds ratio. Univariable analyses and full models in supplementary material S4 and S5 respectively

#### Ward level (CHPPD)

The relationship between staffing measured at postnatal ward level and patient experience responses are displayed in Table 6. There were no statistically significant associations between Registered staffing CHPPD and women’s responses, and the direction of the point estimate was inconsistent in the four questions.

**Table 6:** Summary of univariable and adjusted regression analysis. Estimated odds of a positive response compared to the Lowest tertile of staffing (analysed at Ward level).

|  | Discharge without delay<br>OR(95%CI) |  | Help when needed it<br>OR(95%CI) |  | Information / explanations<br>OR(95%CI) |  | Kindness<br>OR(95%CI) |  |
| --- | --- | --- | --- | --- | --- | --- | --- | --- |
|  | Univariable | Adjusted* | Univariable | Adjusted* | Univariable | Adjusted* | Univariable | Adjusted* |
| **Registered staff CHPPD** |  |  |  |  |  |  |  |  |
| Midtertile | OR 0.98<br>(0.86, 1.23) | OR 0.91<br>(0.78, 1.06) | OR 0.92<br>(0.77, 1.10) | OR 0.76<br>(0.64, 0.91) | OR 0.93<br>(0.80, 1.08) | OR 0.87<br>(0.74, 1.01) | OR 0.98<br>(0.83, 1.15) | OR 0.83<br>(0.69, 1.00) |
| High tertile | OR 1.06<br>(0.93, 1.21) | OR 1.005<br>(0.87, 1.16) | OR 1.05<br>(0.88, 1.25) | OR 0.91<br>(0.77, 1.08) | OR 1.07<br>(0.93, 1.25) | OR 1.01<br>(0.87, 1.18) | OR 1.10<br>(0.93, 1.30) | OR 0.97<br>(0.82, 1.16) |
| **Support worker CHPPD** |  |  |  |  |  |  |  |  |
| Mid tertile | OR 1.02<br>(0.90, 1.17) | OR 1.07<br>(0.93, 1.24) | OR 1.02<br>(0.86, 1.22) | OR 1.12<br>(0.94, 1.32) | OR 0.91<br>(0.78, 1.05) | OR 0.94<br>(0.81, 1.09) | OR 1.06<br>(0.90, 1.25) | OR 1.14<br>(0.96, 1.35) |
| High tertile | OR 1.04<br>(0.91, 1.19) | OR 1.09<br>(0.94, 1.27) | OR 1.14<br>(0.96, 1.36) | OR 1.29<br>(1.08, 1.54) | OR 1.04<br>(0.90, 1.20) | OR 1.09<br>(0.93, 1.27) | OR 1.15<br>(0.97, 1.35) | OR 1.25<br>(1.04, 1.49) |

\* Adjusted for age, parity and type of birth for all models, and number of births per Trust where this improved model fit FTE – full time equivalent, OR odds ratio. Univariable analyses and full models in supplementary material S6 and S7 respectively

Support worker staffing was measured only at ward level and there were differences in the odds of positive responses in Trusts with a higher number of support worker care hours. In the adjusted models, the odds of reporting a positive experience was 25% greater for being treated with kindness and understanding (OR 1.25, 95% CI 1.04,1.49), and 29% greater for reporting having help when needed it (OR 1.29, 95% CI 1.08, 1.54) in the higher staffed Trusts compared to the lowest. Results for the other two questions were in the same direction but not statistically significant.

For overall number of staff (registered plus support staff CHPPD) the adjusted models showed higher odds of reporting positive experiences when the overall number of staff was in the highest compared to lowest tertile, although this was not statistically significant. Trusts in the highest tertile for skill mix (measured as the percentage of Registered staff CHPPD compared to Overall staff CHPPD) had lower odds of a positive response to all questions compared to those in the lowest tertile. This was statistically significant for women reporting they always had help when needing it (OR 0.81, 95% CI 0.70, 0.95) and discharged without delay (OR 0.86, 95% CI 0.76, 0.98) (see supplementary material S8) There was no significant relationship between Obstetric and Gynaecology doctors per 100 births and patient experience. However, this variable was retained in many of the full models as it improved model fit.

#### Sensitivity analyses

We modelled staffing levels as a continuous variable (supplementary material S9) but substantive conclusions were unchanged. The potential for interaction effects was explored by examining the model fit when considering tertiles of Registered staff in combination with different tertiles of health care support staff, and with parity, age and type of birth as categorical variables. No improvement in model fit was noted when these interaction effects were added.

When outliers for CHPPD were removed this resulted in very small changes to the odds ratio estimates and there were no changes to the statistical significance of associations. Effect sizes were smaller when using an alternative dichotomy (no vs yes sometimes, yes always) but the conclusions were unchanged (all sensitivity analyses are presented in supplementary material S10 to S12).

## Discussion

This cross-sectional analysis of linked datasets expands on previous research by using the CHPPD dataset to examine relationships at the ward level using multiple staff groups. We studied the postnatal care experience of women in relation to staffing levels using regression analysis using both Trust level and ward level models. When measured at Trust level, our findings suggest that patient experience is better when more midwives are employed. However, when focussing on postnatal ward staffing measured as Care Hours Per Patient Day, there is no evidence of a relationship between registered midwife/nurse staffing and patient experience. Higher levels of support worker hours were associated with more women reporting they have been treated with kindness and understanding and being helped when they needed it.

Despite having a more granular measurement of staffing at ward level, we have exposed some inconsistencies compared with Trust level staffing which is worthy of further exploration. The weak correlation between FTE midwives and ward level CHPPD is interesting as we expected this to be more closely related. However, this is not an unusual finding as recent research from the USA has also discovered differences in the performance of staffing measures, and called for validated measures to be used[34] Winter et al[23] found that estimates of staffing and outcomes were larger and more precise when data is disaggregated. This has not been found in our study despite the expectation of gaining more clarity with localised data at a ward level.

Our study using the National Maternity Survey data in the preceding year also found a relationship between Trust FTE midwives and better postnatal experience[22]. Although we expected CHPPD to better reflect the staffing experienced by women, CHPPD for registered staff may still be a crude measure of workload as patient acuity and turnover are not factored in. Workload may be variable between postnatal wards with similar staff to patient ratios.

A mismatch may occur between the documented staffing and actual staffing by midwives on postnatal wards, which could result in a measurement bias potentially contributing to dilution or reversal of estimated effects. The deployment models for staff rostered on postnatal wards are not described in the literature. There is evidence that midwives who are rostered to attend postnatal wards are sometimes redeployed during a shift to cover areas of emergent need, such as maintaining one-to-one staffing on labour ward[14, 35, 36]. This has recently been noted in Care Quality Commission reports as the maintenance of one-to-one care in labour resulted in short staffing in other areas because staff were moved at short notice[37, 38]. If these redeployments are not reflected in the roster, units that appear to be highly staffed by midwives may not be.

Our study also examined health care support workers. We found that Trusts in the highest tertile for support worker staffing had more women reporting they always had help when they needed it and were always treated with kindness and understanding. Responses to the questions about delay without discharge and receiving information and explanations were also in the same direction, although not significantly significant when compared with the lowest tertile of support worker staffing. This finding is unsurprising given the nature of the support worker role and the likelihood that they may be available to answer buzzers and contribute to a woman’s experience of feeling supported. An evaluation by Griffin et al[21] reported instances where maternity support workers have facilitated timely discharges, assisted with breastfeeding and parent education. Moreover, midwives were confident in delegating these tasks to the support workers. A survey by Baxter[39] found that women’s satisfaction with postnatal care after caesarean section improved with the introduction of nurses and nursery nurses, highlighting the fact that these roles are valued in this setting. This research on patient experience contrasts with that on clinical outcomes, as Sandall et al[33] reported poorer outcomes for mothers in Trusts where the number of support workers were higher. This composite outcome included some postnatal measures such as length of stay and readmission rates. Overall, this is an under researched area as a recent scoping review found just three studies which related maternity support worker staffing levels with patient outcomes[40]. Our research highlights the importance of examining and adjusting for the contribution of different staff groups. Analysis of results for registered staffing showed differing results before and after adjustment for covariates, including support worker staffing. This study underlines the potential contribution of maternity support workers in the postnatal environment, rather than focusing solely on registered staff. Support workers may be the stable staff on postnatal wards as they are unlikely to be redeployed to care for women arriving in labour.

Our study is limited in that it is cross sectional and we therefore cannot be certain of the precise exposure to staffing levels that each mother was exposed to. One further limitation is that the CHPPD data was identified for postnatal wards in 93 Trusts, and therefore 27 Trusts were not represented in this study. This was because data on CHPPD had not been collected at ward level or we were unable to distinguish postnatal wards from other maternity areas. Although we used a systematic method to identify the postnatal wards, we have not verified this labelling with individual Trusts. We are aware that some wards may have variable activity, such as mixed antenatal and postnatal women, and many include transitional care for high risk babies. In some Trusts the recording staffing appeared to be planned for inpatient maternity services as a whole, and therefore dynamic staffing between areas may be intentional according to patient need. The averaging of staffing data for postnatal wards within the same Trust, and the reporting of CHPPD as monthly averages are sources of imprecision in our analysis, as variability within these measures have not been fully accounted for. Despite some limitations in CHPPD data there is keen interest in this metric as NHS Improvement are considering how patient acuity can be integrated into CHPPD calculations and they are reporting exploratory work linking this measure to patient outcomes[41].

## Conclusion

The relationship between staffing levels and the experience of women on postnatal wards confirms previous research that patient experience is better when more registered midwives are employed in an organisation. The relationship was not seen when registered staffing was measured at a ward level using the Care Hours Per Patient Day dataset, which is a widely used tool for describing staffing levels. An increased number of support workers on postnatal wards was associated with improved maternal experience, highlighting the potential contribution of this sector of the workforce. Further research investigating safety outcomes in relation to postnatal staffing is recommended, as patient experience is one key measure of quality but not the full picture. Limitations of this study mean that a causal relationship cannot be implied and therefore further research is needed to guide policy on postnatal ward staffing.

## Data Availability

The data underlying the analysis cannot be supplied to the journal due to restrictions placed by the UK Data Service https://ukdataservice.ac.uk/app/uploads/cd137-enduserlicence.pdf. The UK Data Service will give access to the data collections only to registered users with a registered use.

