## Supplementary material for "Exploring the relationship between women’s experience of postnatal care and reported staffing measures: an observational study": S1

### S1. Variable selection to go into the model – testing assumption of independence

The following do not appear to be collinear as the VIF is less than 10, therefore safe to go into model together for each of these outcome measures.

Question : DELAY	Question : HELP TIMELY WAY

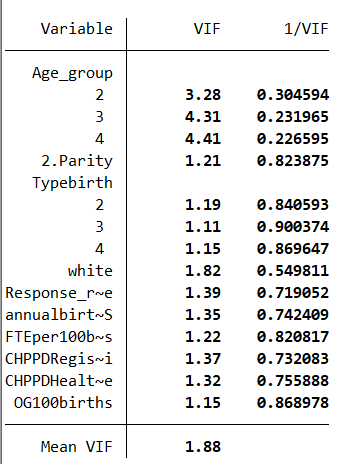

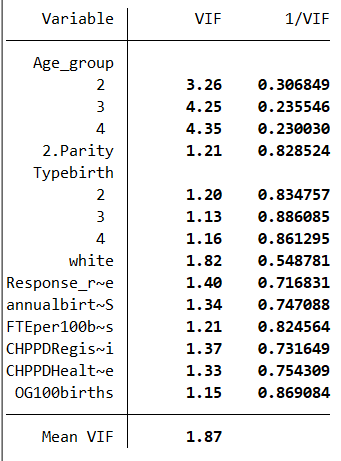

Question : INFO	Question : KINDNESS

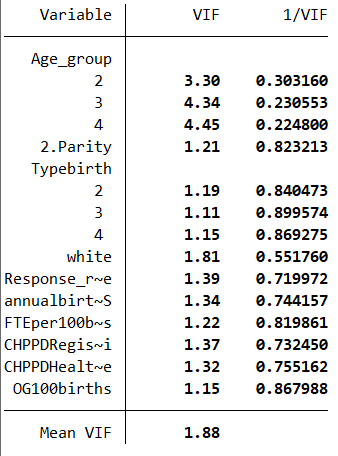

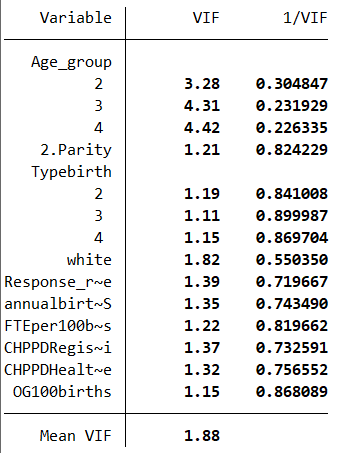

**SELECTING MODELS OF BEST FIT :** Text in red shows model chosen for main analysis

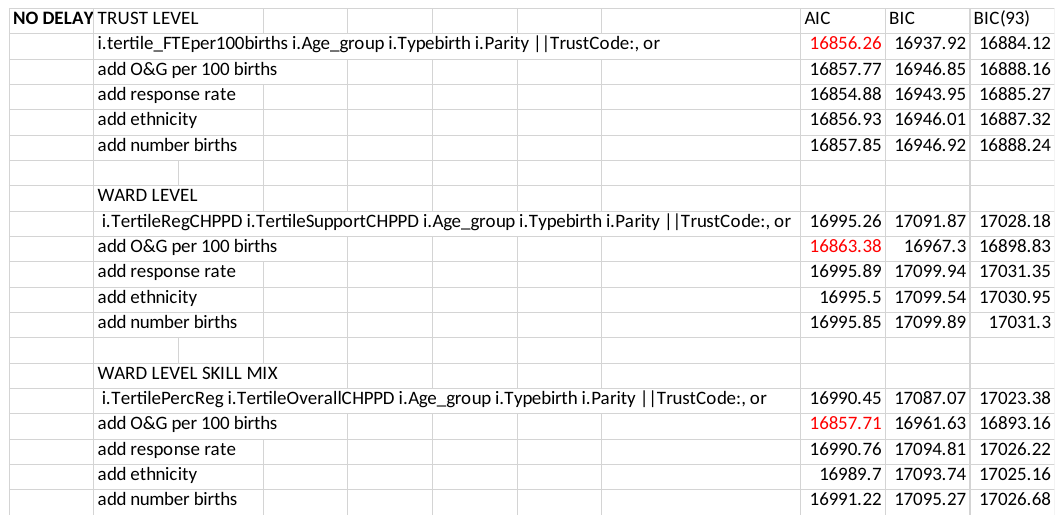

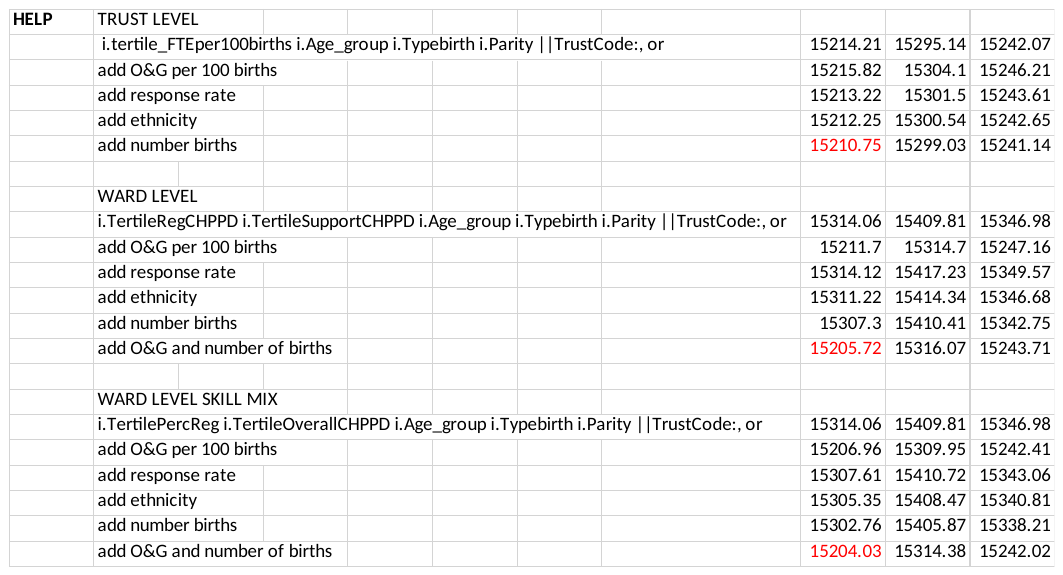

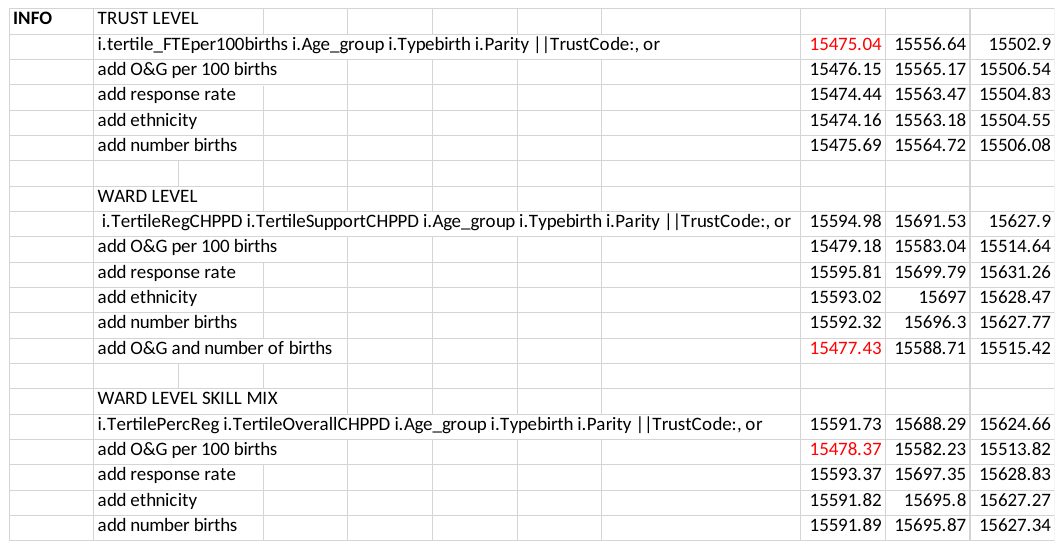

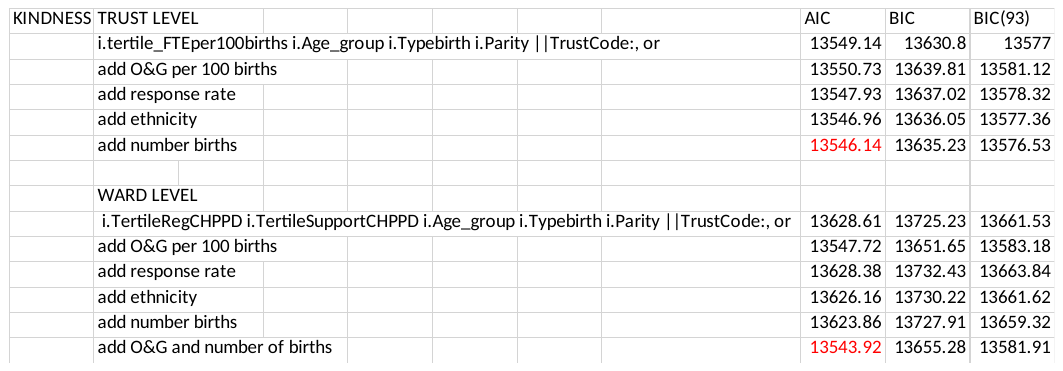
