## Supplementary material for "Exploring the relationship between women’s experience of postnatal care and reported staffing measures: an observational study": S2

### S2. Distribution of staffing data

The patterns of data according to Registered staff and Support staff within Trusts can be seen in Table S1. The shaded areas show better resourced (yellow) and worse resourced Trusts by tertile of staffing (grey)

Table S1 : Distribution of Trusts according to Registered and Support staff by tertile

| Number of Trusts | Low support staff CHPPD tertile  (0.26-2.19) | Mid support staff CHPPD tertile  (2.22-2.91) | HIgh support staff CHPPD tertile  (2.95-6.45) | Total |
| --- | --- | --- | --- | --- |
| Low Registered CHPPD tertile (1.56-4.14) | 21* | 7 | 3 | 31 |
| Mid Registered CHPPD tertile (4.19-5.17) | 5 | 13 | 13 | 31 |
| High Registered CHPPD tertile  (5.24-9.57) | 6 | 10 | 15 | 31 |
| Total | 32 | 30 | 31 | 93 |

*two Trusts on the border had identical values for support worker staffing, therefore 21 in this category

Trust level(y) and ward level (x) measures of staffing appear to have little relationship between them (on visual inspection).

Figure S1 : Scatterplot of Registered staff CHPPD and FTE midwives per organisation

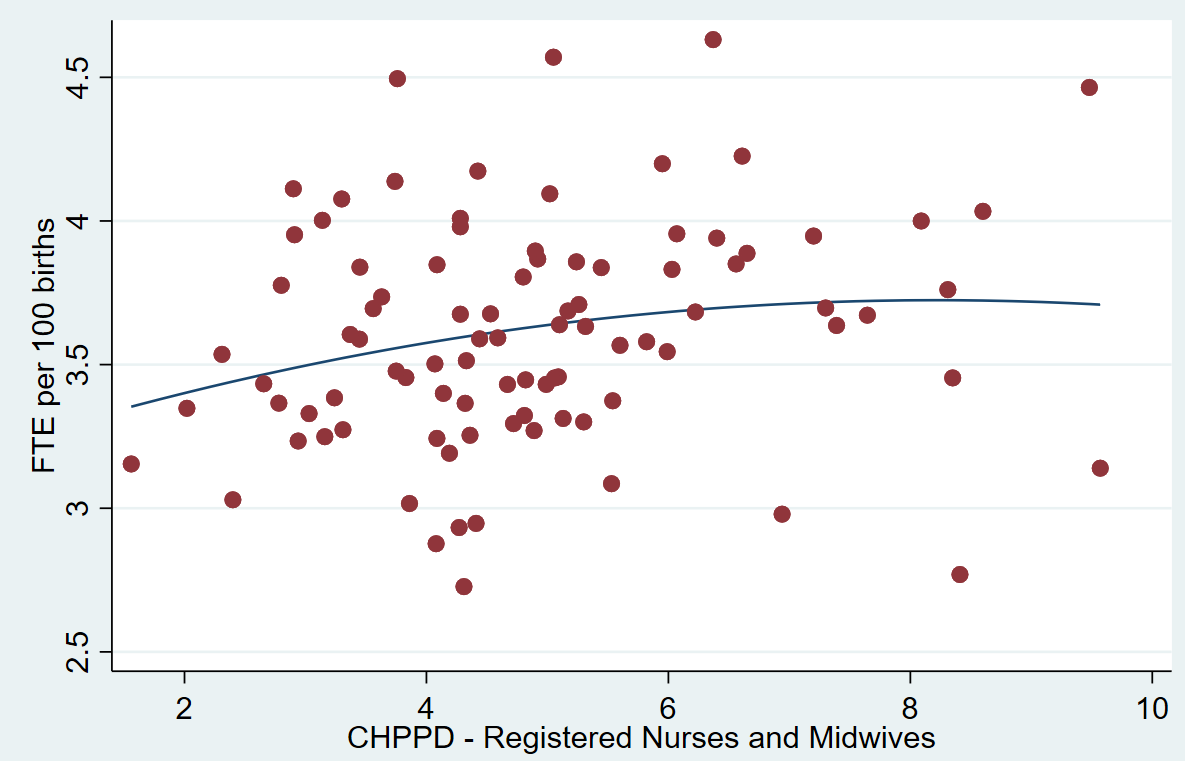

A weak relationship is seen when Pearson and Spearmans coefficients are calculated.

Pearson Correlation coefficient between these two variables is 0.1388, p=0.000
This measures of the strength of a linear association between two variables.
Spearman's rho =	0.1974	Nonparametric test of independence, Prob > |t| =	0.0000
