## Supplementary material for "Exploring the relationship between women’s experience of postnatal care and reported staffing measures: an observational study": S3

### S3. Univariable analyses for age group, parity, type of birth, ethnicity, number of births in the Trust, response rate and medical staff

**Question related to being Discharged without delay**

|  |  | Indication of model fit |
| --- | --- | --- |
|  | Empty model | AIC 17558.92 |
| Age group (comparison <25)  25 - 29 year olds 30 - 34 year olds  35+ year olds | 1.18 (1.01, 1.38) 1.26 (1.09, 1.46) 1.30 (1.12, 1.51) | AIC 17551 |
| Parity (comparison primiparous) Multiparous | 1.43 (1.33, 1.53) | AIC 17228.36 |
| Type birth  (comparison spontaneous birth)  instrumental birth planned caesarean emergency caesarean | 0.67 (0.61, 0.74) 1.08 (0.97, 1.20) 0.87 (0.78, 0.96) | AIC 17259.94 |
| % white ethnicity | 1.00 (1.00, 1.00) | AIC 17559.11 |
| Number of births in Trust per year | 1.00 (1.00, 1.00) | AIC 17559.5 |
| % response rate per Trust | 1.00 (1.00, 1.01) | AIC 17559.71 |
| FTE O&Gper100births | 1.00 (0.77, 1.31) | AIC 17424.37 |

**Question related to Always having help when needed it**

|  |  | Indication of model fit |
| --- | --- | --- |
|  | Empty model | AIC 15864.99 |
| Age group (comparison <25)  25 - 29 year olds 30 - 34 year olds  35+ year olds | 1.071 (.908, 1.264) 1.049 (.897, 1.226) 1.053 (.899, 1.232) | AIC 15870.32 |
| Parity (comparison primiparous) Multiparous | .701 (.653, .753) | AIC 15588.74 |
| Type birth  (comparison spontaneous birth)  instrumental birth planned caesarean emergency caesarean | .649 (.583, .723) .691 (.619, .771) .680 (.613, .754) | AIC 15536.37 |
| % white ethnicity | 1.006 (1.001, 1.010) | AIC 15862.09 |
| Number of births in Trust per year | 1.000 (1.000, 1.000) | AIC 15859.81 |
| % response rate per Trust | 1.007 (.997, 1.017) | AIC 15865.23 |
| FTE O&Gper100births | .950 (.670, 1.347) | AIC 15766.32 |

**Question related to Always having Info and explanations**

|  |  | Indication of model fit |
| --- | --- | --- |
|  | Empty model | AIC 16374.35 |
| Age group (comparison <25)  25 - 29 year olds 30 - 34 year olds  35+ year olds | .967 (.820, 1.140) .993 (.849, 1.160) 1.08 (.921, 1.262) | AIC 16374.9 |
| Parity (comparison primiparous) Multiparous | 1.823 (1.691, 1.967) | AIC 15890.25 |
| Type birth  (comparison spontaneous birth)  instrumental birth planned caesarean emergency caesarean | .527 (.475, .587) .738 (.660, .825) .548 (.495, .607) | AIC 15954.53 |
| % white ethnicity | 1.004 (1.000, 1.008) | AIC 16372.26 |
| Number of births in Trust per year | 1.000 (1.000, 1.000) | AIC 16372.45 |
| % response rate per Trust | 1.004 (.995, 1.012) | AIC 16375.57 |
| FTE O&Gper100births | .947 (.706, 1.271) | AIC 16267.14 |

**Question related to Always being treated kindness and understanding**

|  |  | Indication of model fit |
| --- | --- | --- |
|  | Empty model | AIC 14288.45 |
| Age group (comparison <25)  25 - 29 year olds 30 - 34 year olds  35+ year olds | 1.363 (1.144, 1.622) 1.319 (1.120, 1.553) 1.373 (1.164, 1.620) | AIC 14279.93 |
| Parity (comparison primiparous) Multiparous | 1.731 (1.592, 1.881) | AIC 13916.3 |
| Type birth  (comparison spontaneous birth)  instrumental birth planned caesarean emergency caesarean | .561 (.500, .631) .627 (.557, .707) .512 (.458, .571) | AIC 13920.35 |
| % white ethnicity | 1.006 (1.002, 1.011) | AIC 14282.91 |
| Number of births in Trust per year | 1.000 (1.000, 1.000) | AIC 14283.7 |
| % response rate per Trust | 1.008 (.999, 1.018) | AIC 14287.22 |
| FTE O&Gper100births | .893 (.639, 1.247) | AIC 14210.48 |
