## Supplementary material for "Exploring the relationship between women’s experience of postnatal care and reported staffing measures: an observational study": S4

### S4. Relationship between staffing and patient experience at Trust level (univariable)

Mid and Highest tertile compared with lowest tertile as the reference group

**Question related to being Discharged without delay**

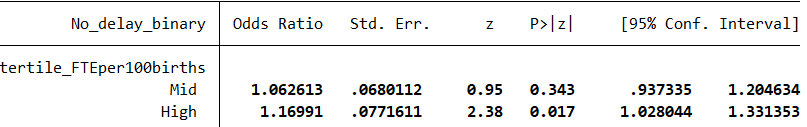

**Question related to Always having help when needed it**

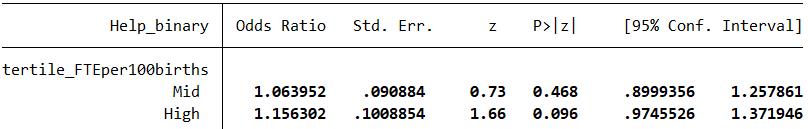

**Question related to Always having Info and explanations**

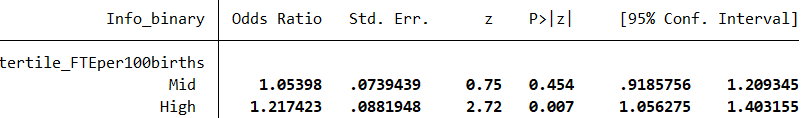

**Question related to Always being treated kindness and understanding**

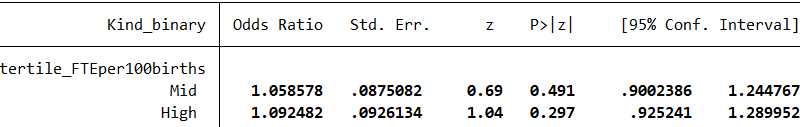
