## Supplementary material for "Exploring the relationship between women’s experience of postnatal care and reported staffing measures: an observational study": S5

### S5. Relationship between staffing and patient experience at Trust level (adjusted models based on model of best fit by AIC)

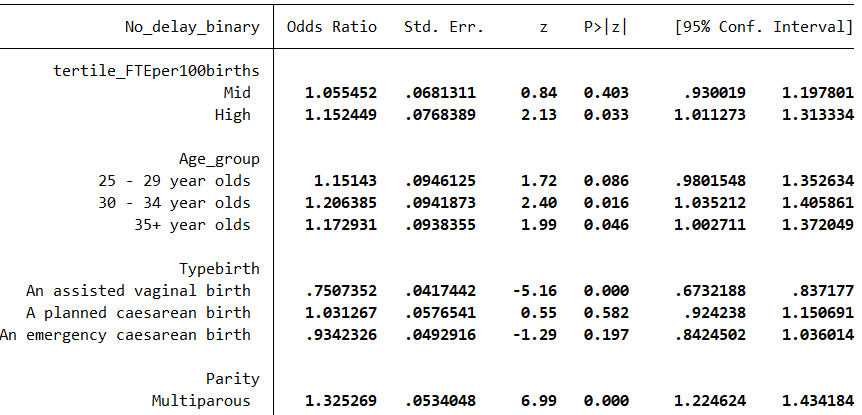

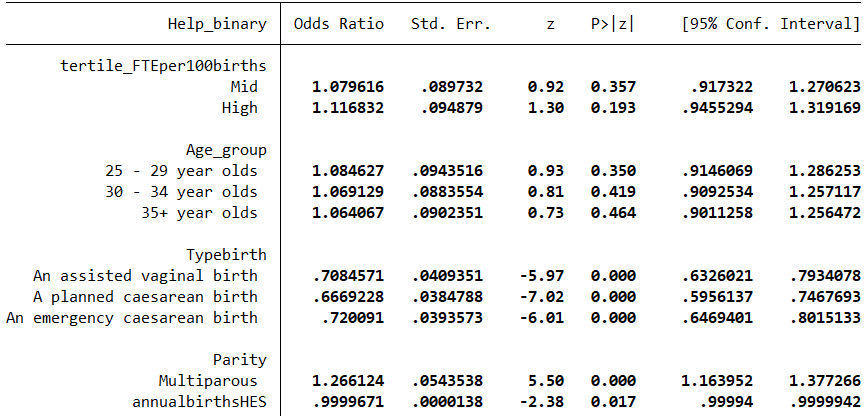

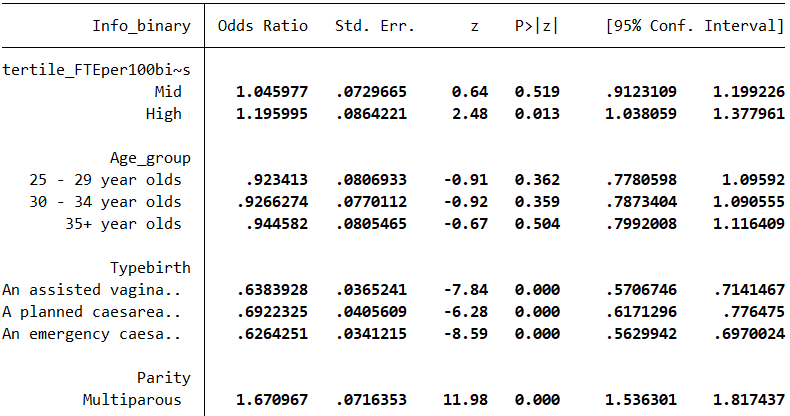

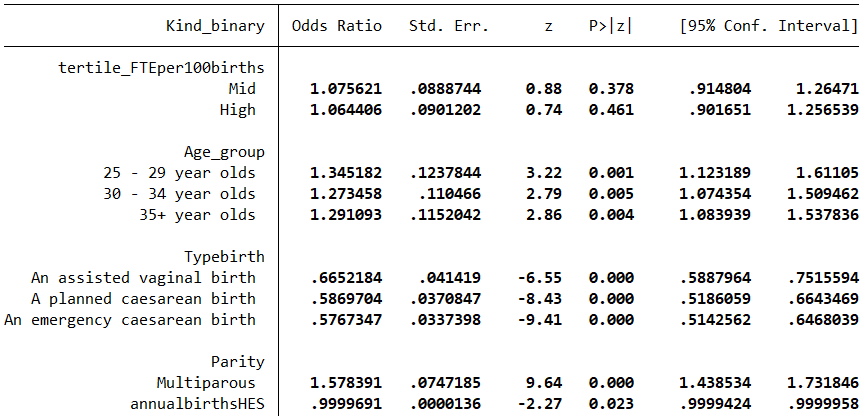
