## Supplementary material for "Exploring the relationship between women’s experience of postnatal care and reported staffing measures: an observational study": S6

### S6. Relationship between staffing and patient experience at postnatal ward level (univariable)

The tables below show results for tertiles of Registered staff CHPPD and Support staff CHPPD measured at postnatal ward level for each of the 4 questions. The analyses are in relation to the tertile with the lowest staffing.

**Question related to being Discharged without delay**

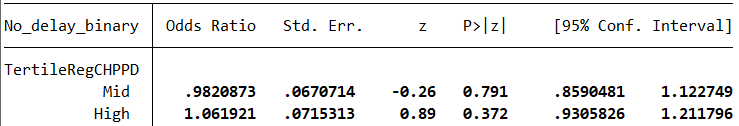

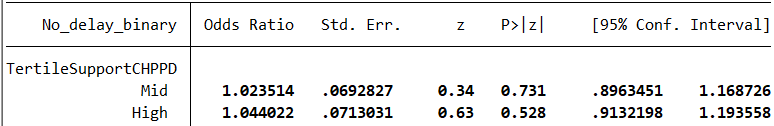
 **Question related to Always having help when needed it**

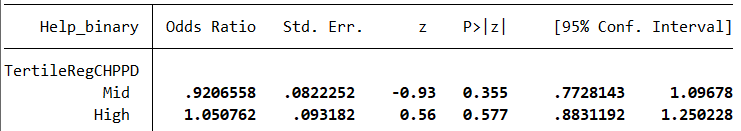

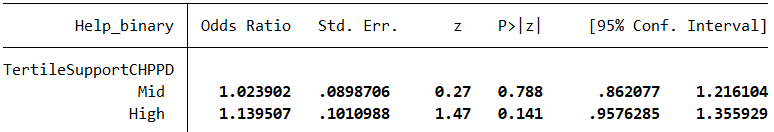

**Question related to Always having Info and explanations**

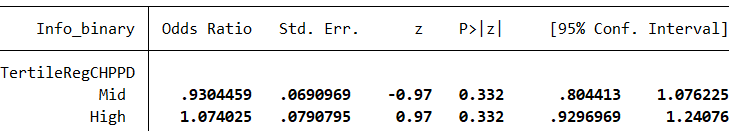

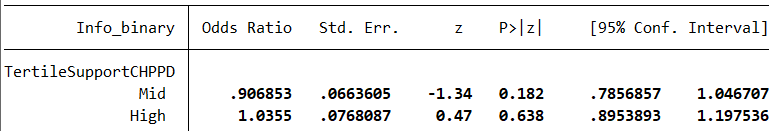

**Question related to Always being treated kindness and understanding**

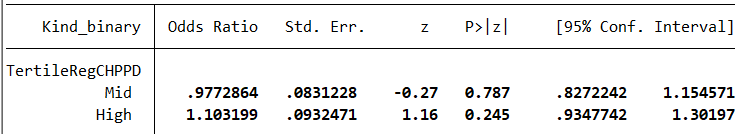

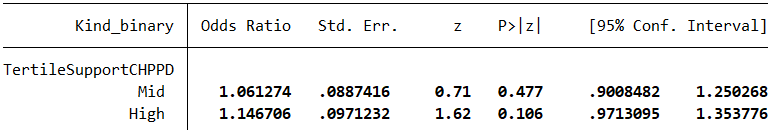
