## Supplementary material for "Exploring the relationship between women’s experience of postnatal care and reported staffing measures: an observational study": S8

### S8. Relationship between Overall staffing, Skill mix and patient experience at postnatal ward level (adjusted models, which were best fit according to AIC)

**Question related to being Discharged without delay**

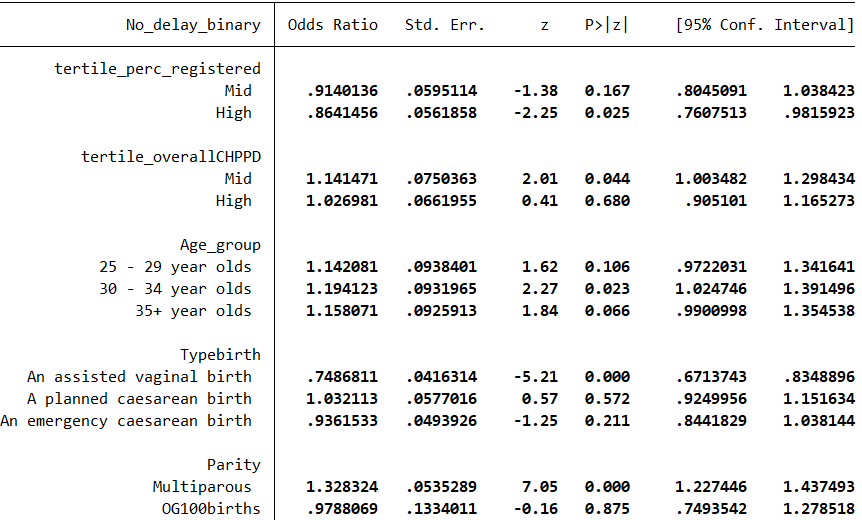

**Question related to Always having help when needed it**

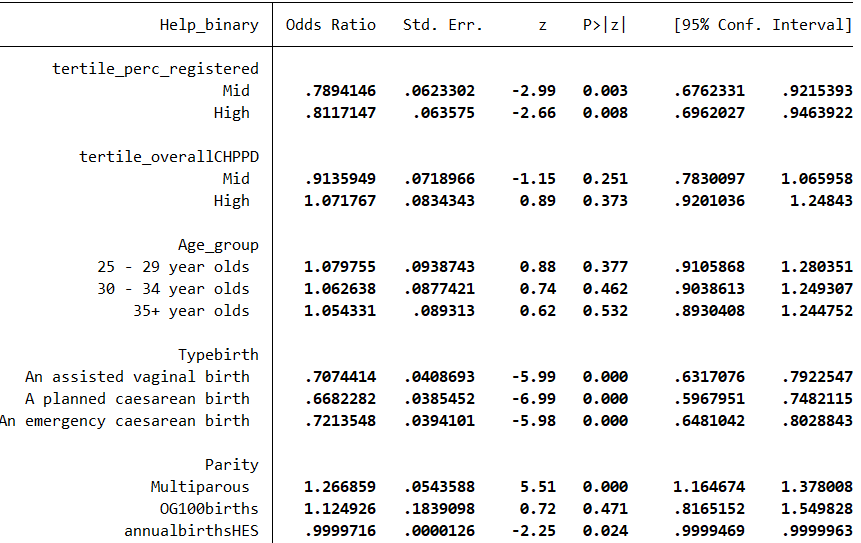

**Question related to Always having Info and explanations**

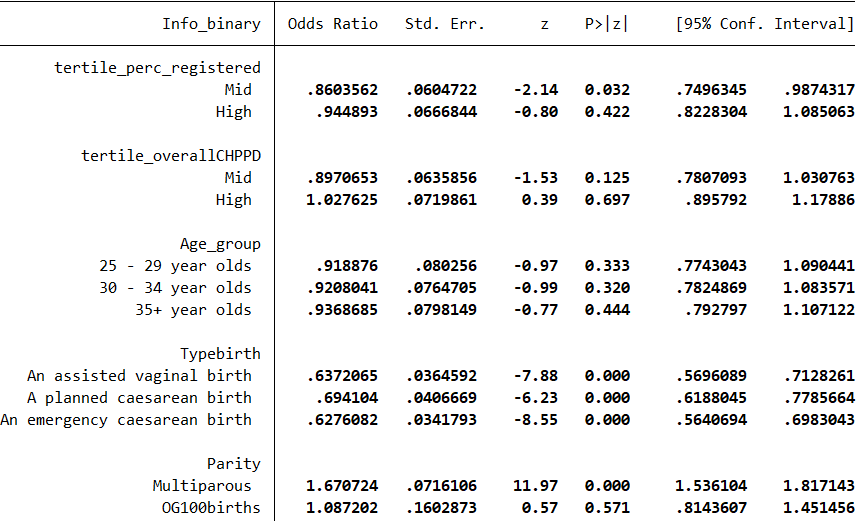

**Question related to Always being treated kindness and understanding**

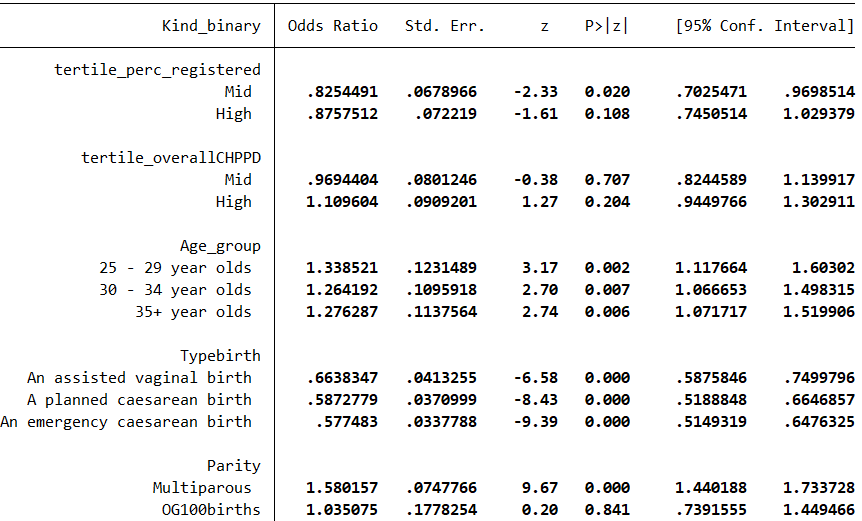
