## Supplementary material for "Exploring the relationship between women’s experience of postnatal care and reported staffing measures: an observational study": S9

### S9. Sensitivity analysis : secondary analyses with staffing as a continuous variable (not tertiles). Extracts from adjusted models presented

TRUST LEVEL

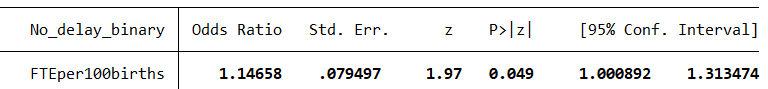

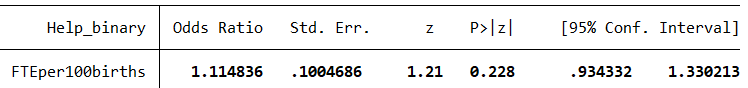

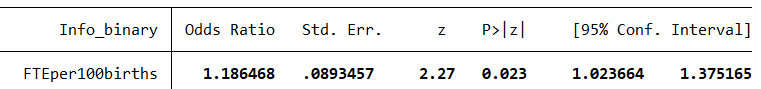

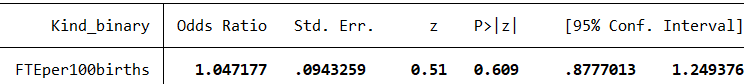

WARD LEVEL

Staff groups

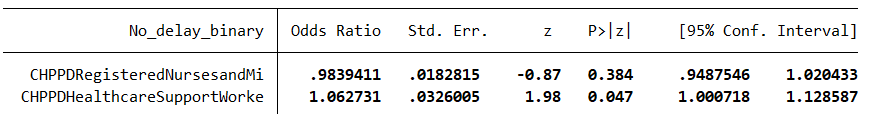

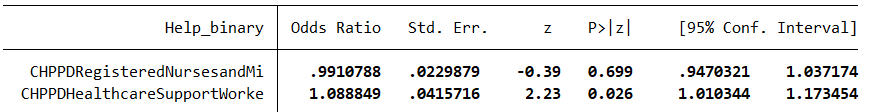

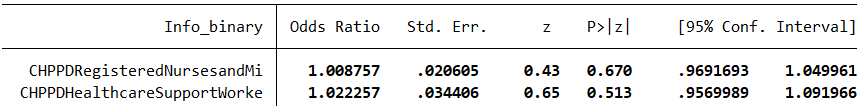

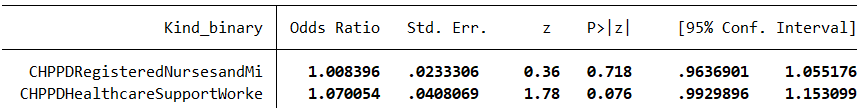

Overall staff and skill mix

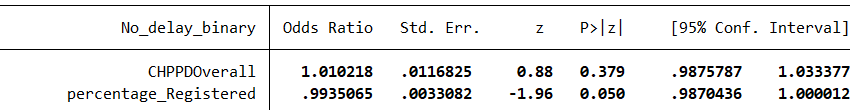

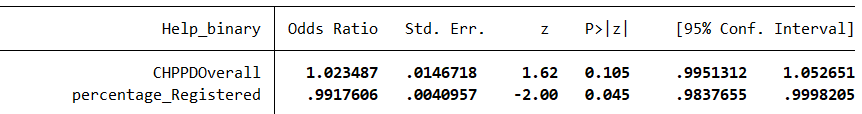

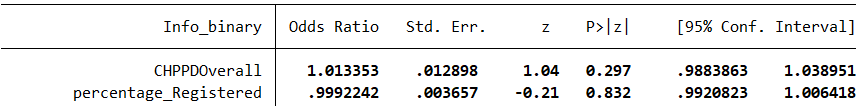

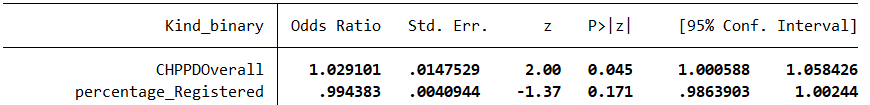
