## Supplementary material for "Exploring the relationship between women’s experience of postnatal care and reported staffing measures: an observational study": S10

### S10. Sensitivity analysis : Testing model fit using AIC and BIC when including interaction variables

|  |  | | **Best fit?** |
| --- | --- | --- | --- |
| Delay in discharge | With no interaction variables | AIC 16863.38  BIC (93) 16898.83 BIC 16967.3 | Reference |
|  | With interaction variable RegisteredxHCSW | AIC 16863.04 BIC (93) 16908.63 BIC 16996.66 | Best fit without interaction included |
|  | With interaction variable RegisteredxParity | AIC 16864.22 BIC (93) 16904.74 BIC 16982.99 | Best fit withoutinteraction included |
|  | With interaction variable RegisteredxType birth | AIC 16873.48 BIC (93) 16924.14 BIC 17021.95 | Best fit without interaction included |
|  | With interaction variable RegisteredxAge group | AIC 16872.84 BIC (93) 16923.5 BIC 17021.31 | Best fit without interaction included |
| Help in reasonable time | With no interaction variables | AIC 15205.72  BIC (93) 15243.71 BIC 15316.07 | Reference |
|  | With interaction variable RegisteredxHCSW | AIC 15207.71 BIC (93) 15255.83 BIC 15347.49 | Best fit without interaction included |
|  | With interaction variable RegisteredxParity | AIC 15207.09 BIC (93) 15250.14 BIC 15332.16 | Best fit without interaction included |
|  | With interaction variable RegisteredxType birth | AIC 15214.44 BIC (93) 15267.63 BIC 15368.94 | Best fit without interaction included |
|  | With interaction variable RegisteredxAge group | AIC 15216.32 BIC (93) 15269.5 BIC 15370.82 | Best fit without interaction included |
| Information/Explanations | With no interaction variables | AIC 15477.43  BIC (93) 15515.42 BIC 15588.71 | Reference |
|  | With interaction variable RegisteredxHCSW | AIC 15483.32 BIC (93) 15531.44 BIC 15624.28 | Best fit without interaction included |
|  | With interaction variable RegisteredxParity | AIC 15480.26 BIC (93) 15523.31 BIC 15606.38 | Best fit without interaction included |
|  | With interaction variable RegisteredxType birth | AIC 15483.24 BIC (93) 15536.43 BIC 15639.03 | Best fit without interaction included |
|  | With interaction variable RegisteredxAge group | AIC 15483.26 BIC (93) 15536.45 BIC 15639.05 | Best fit without interaction included |
| Treated with kindness and understanding | With no interaction variables | AIC 13543.92  BIC (93) 13581.91 BIC 13655.28 | Reference |
|  | With interaction variable RegisteredxHCSW | AIC 13549.45 BIC (93) 13597.57 BIC 13690.5 | Best fit without interaction included |
|  | With interaction variable RegisteredxParity | AIC 13546.39 BIC (93) 13589.44 BIC 13672.59 | Best fit without interaction included |
|  | With interaction variable RegisteredxType birth | AIC 13550.38 BIC (93) 13603.56 BIC 13706.28 | Best fit without interaction included |
|  | With interaction variable RegisteredxAge group | AIC 13551.62 BIC (93) 13604.8 BIC 13707.52 | Best fit without interaction included |
