## Supplementary material for "Exploring the relationship between women’s experience of postnatal care and reported staffing measures: an observational study": S11

### S11. Sensitivity analysis after removing outliers

Mild outliers for CHPPD were removed when they were more than 1.5 x IQR more unusual than Q1 or Q3. This resulted in the removal of data for 2 Trusts for Registered CHPPD data, no outlying Trusts for Support CHPPD data, and 1 Trust for Overall CHPPD data. There were no Extreme outliers classified as 3 x IQR more unusual than Q1 or Q3

Method of identifying outliers taken from Dunn P. Scientific Research and Methodology : An introduction to quantitative research and statistics in science, engineering and health 2021.<https://bookdown.org/pkaldunn/Book/identifying-outliers.html>

Removing the Trusts meant that individual patient data was removed from the analysis and the remaining Trusts were grouped into different tertiles of CHPPD which altered their analysis.

The new tertiles were
CHPPD Registered staff	1.56 to 4.09 (low), 4.14 to 5.10 (medium) and 5.13 to 8.6 (high)
CHPPD Overall staff 1.82 to 6.4 (low), 6.59 to 8.2 (medium) and 8.36 to 12.91 (high)

The models in part G (above) have been repeated using data with outlying values removed and treated as missing.

**Question related to being Discharged without delay**

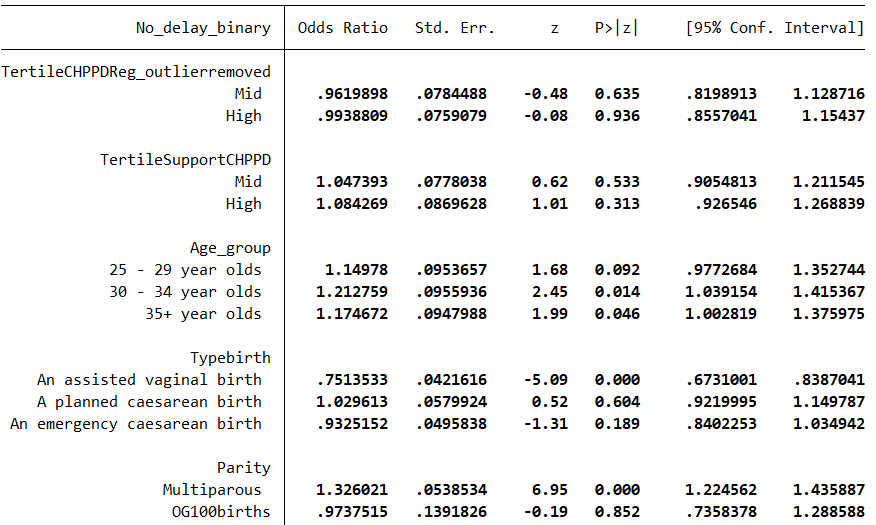

**Question related to Always having help when needed it**

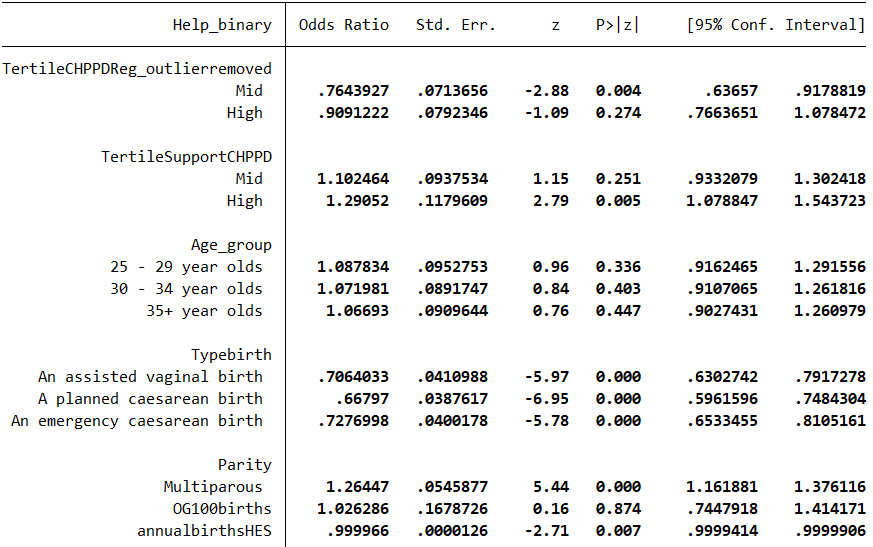

**Question related to Always having Info and explanations**

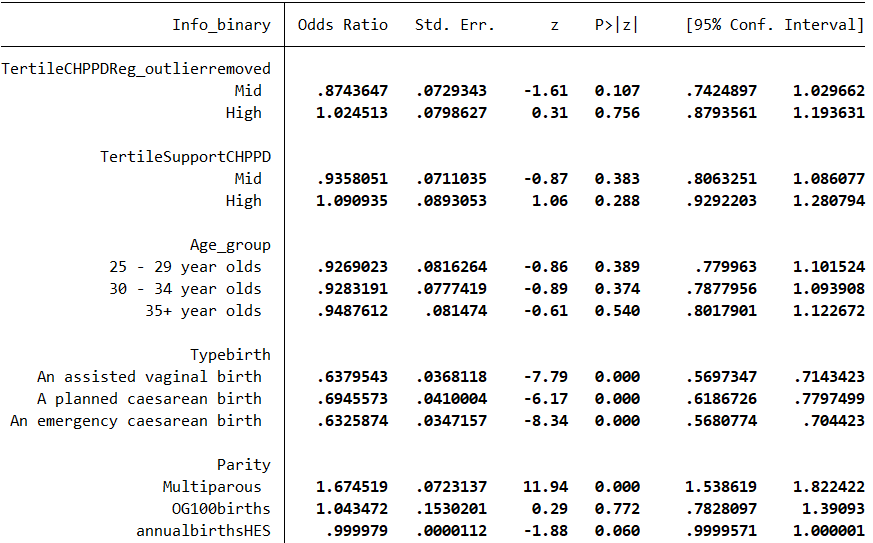

**Question related to Always being treated kindness and understanding**

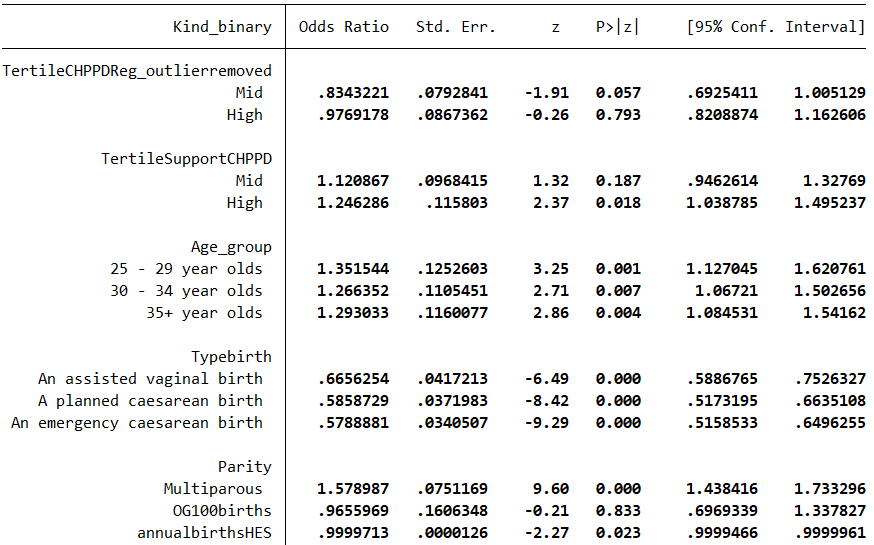
