## Supplementary material for "Exploring the relationship between women’s experience of postnatal care and reported staffing measures: an observational study": S12

### S12. Sensitivity analyses using alternative dichotomy of question responses

The models in part G (above) have been repeated using data with recoding of responses,
with yes always / yes sometimes = 1, no=0

**Help when you needed it?**

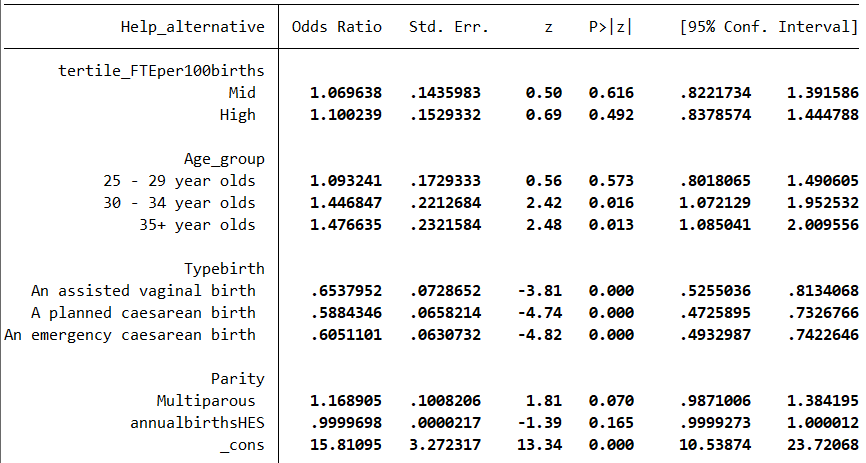

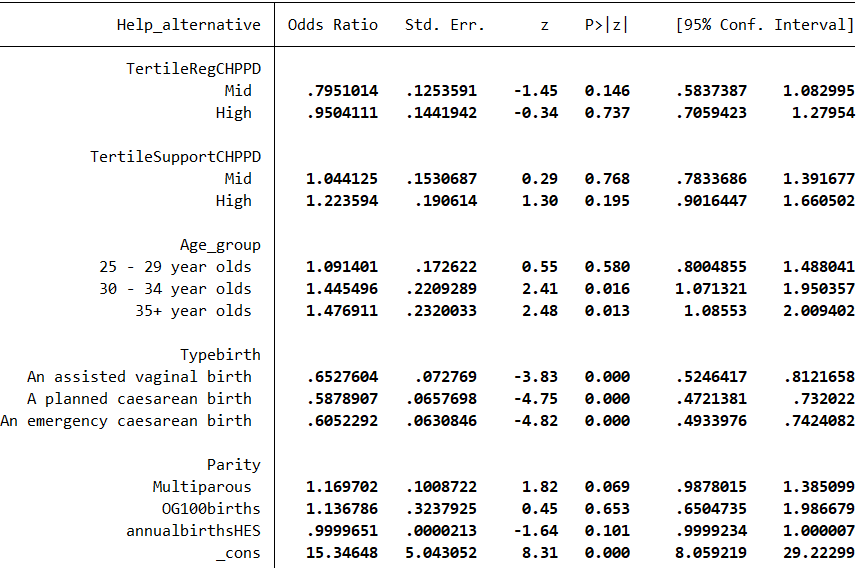

**Given the information or explanations you needed?**

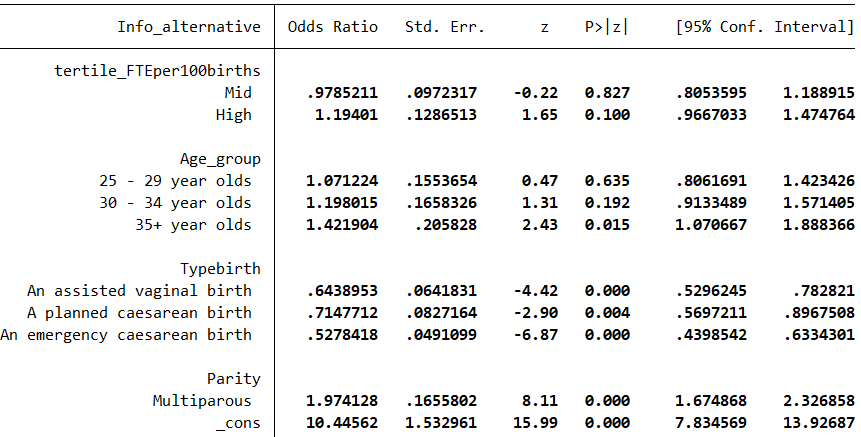

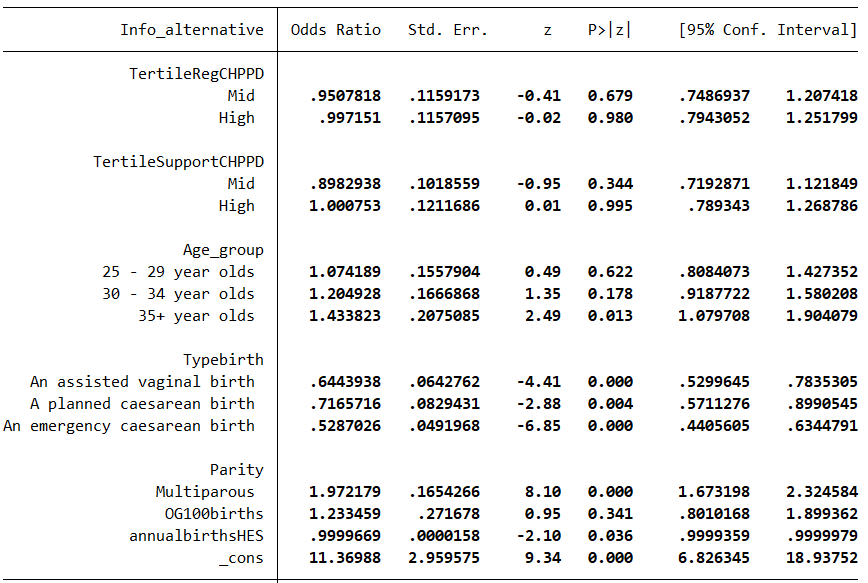

**Treated with kindness and understanding?**

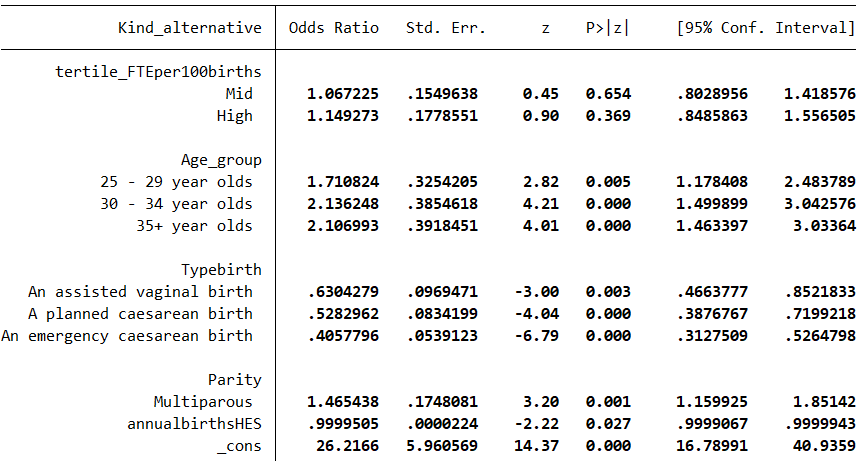

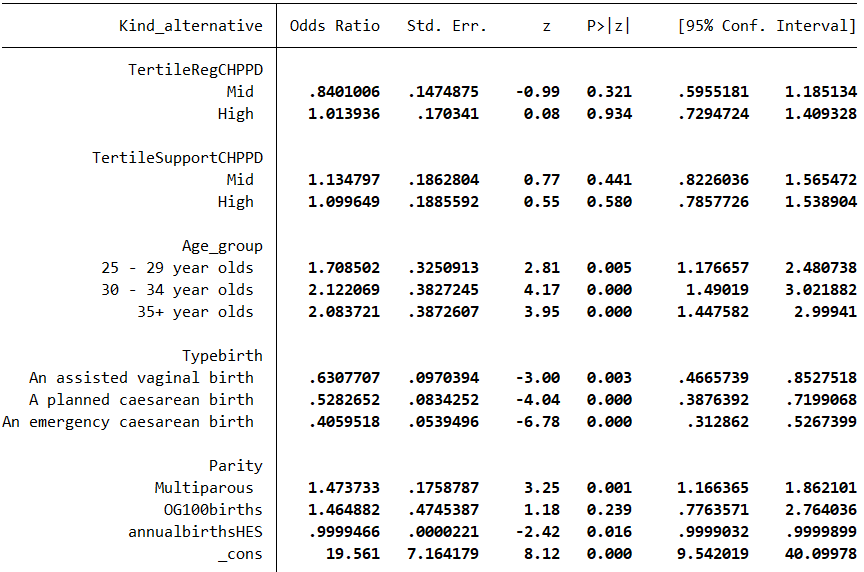
